# A tool for mapping medical narratives into medical ontologies in low resource settings: A case study for German

**DOI:** 10.1101/2024.06.11.24307163

**Authors:** Faizan E Mustafa, Juan G. Diaz Ochoa

## Abstract

Named Entity Recognition (NER) is extremely relevant in the clinical field since it allows the extraction of information, such as diagnoses or medical procedures, from non-structured data (doctor’s letters, vignettes, etc.) and coding them based on international classification systems. As a result, language models should be trained to recognize and classify these items accurately. While Large Language Models (LLMs) like ChatGPT are capable of recognizing medical entities in texts, they are not reliable at performing this task. Unlike English, where there are a variety of resources to assist with this task, other languages, such as German, lack appropriate language models. This study presents a methodology for the generation of high-quality full-synthetic datasets and the implementation of a workflow for the identification and classification of diseases, co-diseases, and medical procedures for clinical narratives in oncology.

## Introduction

In health institutions, the extraction of information from unstructured healthcare data and clinical narratives is critical at the clinical and administrative levels (for example, to determine medical procedures or to obtain relevant information for reimbursement purposes). As an example, diseases can be coded (using the International Disease Classification ICD) using Natural Language Processing (NLP).

To this end, Large Language Models (LLM) can be used to perform this task. However, recent investigations have shown that LLMs are poor coders [1]. Furthermore, LLMs are usually deployed as cloud applications, which are essentially non-conform with data protection policies required to deal with health data. Thus, models that can be implemented at the edge and performing with high precision are required.

Currently there is restricted availability of corpora and annotated data for NLP in medicine in languages other than English. We implemented a pipeline based on the generation of synthetic narratives for named entity recognition model training (to identify diseases and procedures in medical texts) and entity linking to link recognized entities to ICD and OPS (medical procedure) classifications.

We deployed the trained models as a tool to assist customers identify relevant items in clinical narratives for reimbursement. The implementation has been performed in German but can be extrapolated into other languages.

## Methods and Data

Real narratives are problematic from a data protection perspective. This is particularly critical in the development of AI models outside hospitals. Even if the model is deployed without a database, there is a probability that with reverse engineering critical data can be extracted from the model [2].

We create a synthetic dataset to train NER model without using any real patient data following the method used for the creation of the Aluminium standard [3]. We extracted medical entities (main disease, co-disease, medication, procedure) from the original narratives that are then automatically introduced in a prompt to generate the synthetic narrative through ChatGPT. The created synthetic narratives were manually annotated using Doccano and used to train NER model with SCAI-BIO/bio-gottbert-base [4] as base model for 20 epochs. We use SapBERT model finetuned on German biomedical data [5] to link the recognized entities by the NER model to ICD-10 (for diseases) and OPS ontology (for medical procedures). Finally, the trained model is deployed using PyDash on MS-Azure.

## Results

NER validation results are reported for “Type” evaluation scheme [2] in Table 1. Since diagnoses usually have larger spans, they are more prone to be context dependent than procedures or medications (which are often provided in the medical narratives as single words or spans containing 2 to 3 words). Thus, validation values differ, and are often better for medical procedures and medications (with f1 > 0.5) and poorer for co-diagnoses (with f1 < 0.5).

**Table 1.**
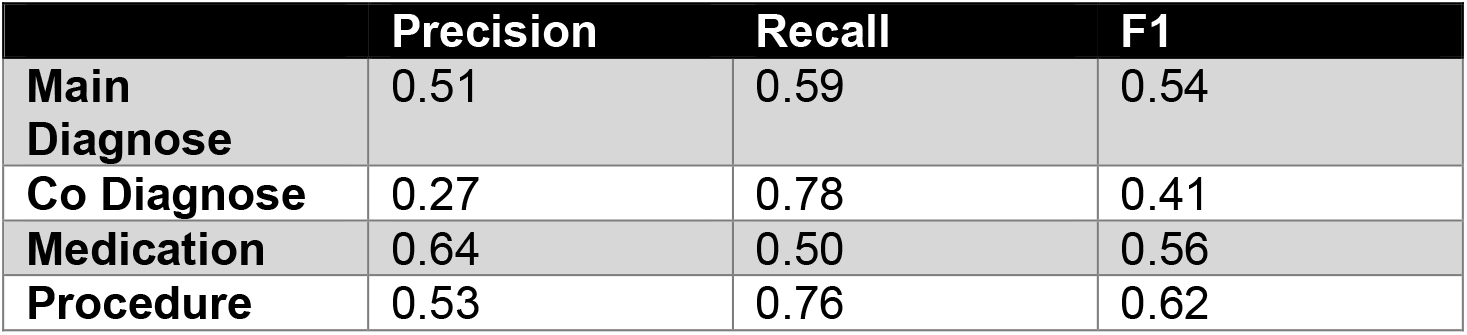
NER evaluation results.

|  | Precision | Recall | F1 |
| --- | --- | --- | --- |
| **Main Diagnose** | 0.51 | 0.59 | 0.54 |
| **Co Diagnose** | 0.27 | 0.78 | 0.41 |
| **Medication** | 0.64 | 0.50 | 0.56 |
| **Procedure** | 0.53 | 0.76 | 0.62 |

We have developed a demonstrator that identifies diagnostic and procedural elements within clinical narratives and links them to ontology entities. It occurs after a clinical narrative has been entered (see Fig. 1). The NER-recognized entities are highlighted in the text, along with their associated ontology entities. The customer can also select an appropriate ICD or OPS entity from the dashboard. This pipeline is easily adaptable to languages with limited resources.

**Figure 1.**
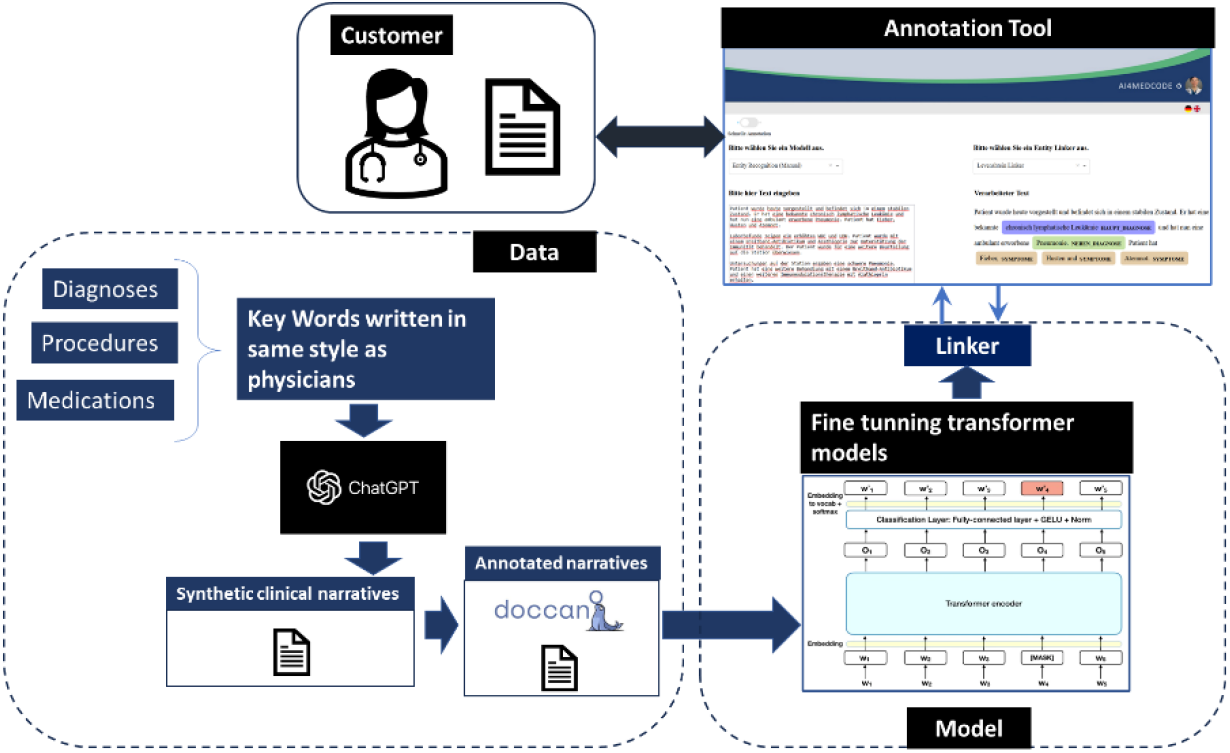
A pipeline for training the demonstrator models. In the diagram, a snapshot of the implemented solution is displayed, where the annotator (customer) can scan her documents and extract classifications (ICD for Diagnoses and OPS for medical procedures).

## Conclusion

We are presenting a demonstrator for a semi-automatic annotation tool for clinical narratives. In order to protect patient data, training data has been fully synthesized, in what we call a synthetic gold standard. The current implementation does not exclude the possibility to further fine tune the model using a silver standard based on bio-medical terminology (as reported by Wang et al. [6]). The method presented here allows us to train and deploy tools for text annotation in any language while protecting the privacy of original patient data. We plan to synthesize narratives using off-the-shell models like LLama2/3 / Mistral in future implementations.

### Study context

This project was supported by the Ministry for Economics, Labor and Tourism from Baden-Württemberg, Germany via grant agreement number BW1_1456 (AI4MedCode).

## Data Availability

All data produced in the present study, which is fully synthetic, are available upon reasonable request to the authors

### Appendix: Methodology

The keywords for the generation of the synthetic narratives have been edited by a physician, figuring out which are the typical combination of main diseases, co-diseases, symptoms and medical procedures. Such keywords are then introduced in a pipeline in a text-generation program. We used ChatGPT as well as Mistral for text generation.

Different synthetic narratives are generated for different patients and stored in different documents, similar to real clinical documents. Thereafter the narratives were manually annotated using Doccano. We preferred a manual annotation, first because we have designed our own annotation rules and, second, because automatic annotation is not reliable enough^1^.

The model was finally trained using the SCAI-BIO/bio-gottbert-base model. The model is finetuned on the train data for 20 epochs. At the end of the finetuning, the best-performing model (best f1 score) on the validation set is selected to get the results for the test set.

Some letters are longer than 512 sub-word tokens which exceeds the limit of the transformer model therefore we split a sample letter into segments of max 4 sentences if the number of subwords tokens is more than 300. The number of sub-words is checked by tokenizing a sample with “Bert-base-German-cased” tokenizer from Huggingface library. The sentence splitting is done using the “de_core_news_sm” model from the SPACY library.

The following are the validation results by entity (diagnose, procedure or medicament) considering different matchings (see Table A-1).

**Table A-1.**
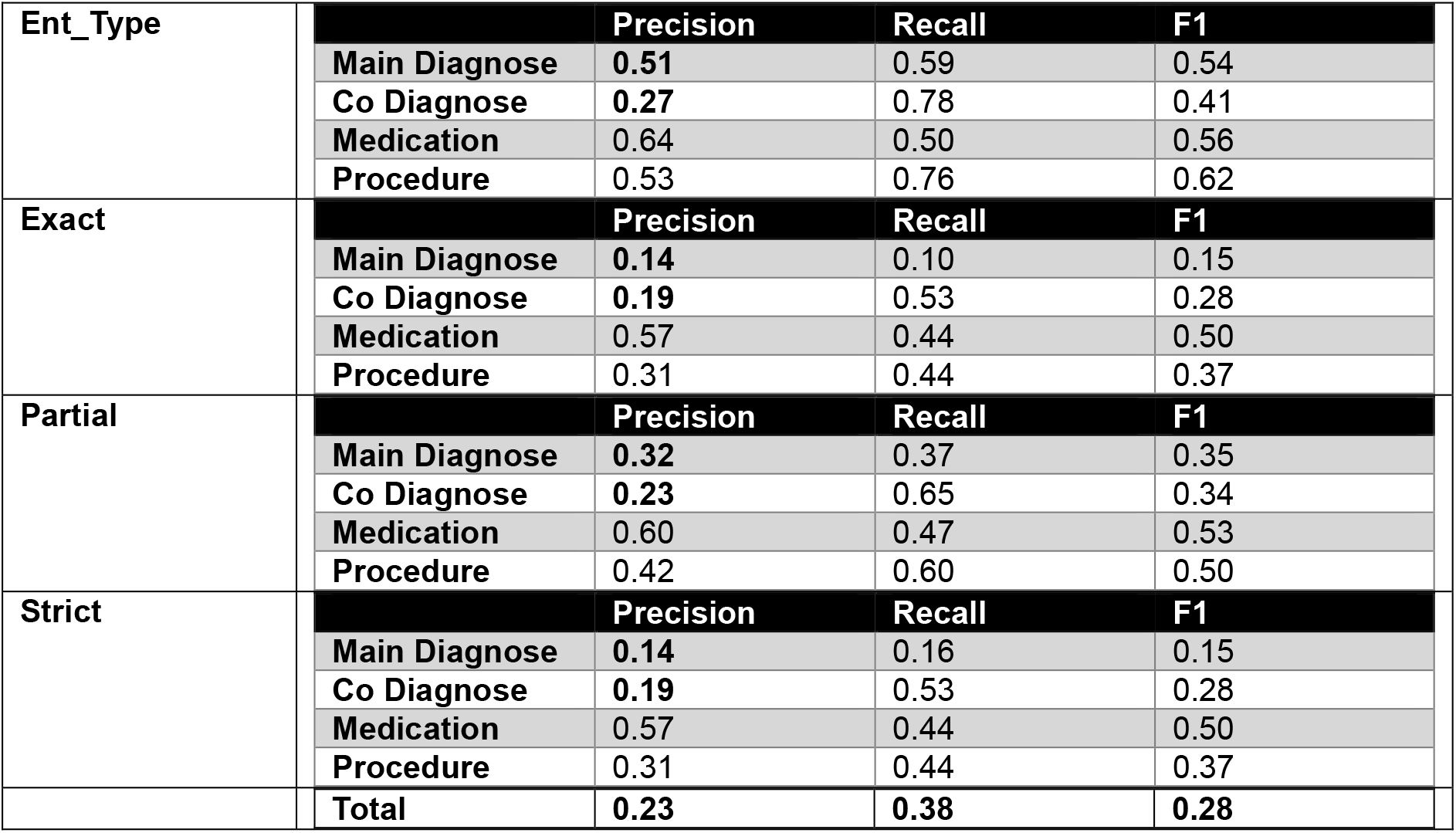
Full validation results considering entity-type, exact, partial and strict matching.

In table 1 in the main article, we only report validation results for entity type.

## Footnotes

1 https://ai.nejm.org/doi/full/10.1056/AIdbp2300040

